# Estimating COVID-19 contribution to total excess mortality

**DOI:** 10.1101/2020.09.10.20191965

**Authors:** Ville N. Pimenoff, Miriam Elfström, Iacopo Baussano, Mikael Björnstedt, Joakim Dillner

## Abstract

We compared the total excess mortality per week in relation to the reported Covid-19 related deaths in the Stockholm region (Sweden). Total excess mortality peaked under the weeks of high COVID-19-related mortality, but 25% of these deaths were not recognized as Covid-related. Most of these deaths occurred outside hospitals. Total all-cause mortality in excess to average all-cause mortality during the epidemic peak period may provide a comprehensive picture of the total burden of COVID19-related deaths.

## Main text

The large number of COVID-19-related deaths reported globally (>850,000 deaths)^1^ may be difficult to compare because of differences in national reporting systems. We aimed to investigate how a readily standardised measure, the excess mortality, was related to the reported COVID-19 deaths in a region with comprehensive health data registries that had been hit by the COVID-19 epidemic, the Stockholm region in Sweden.

We retrieved mortality data for 2020 from the Swedish National Board of Health and Welfare and mortality estimates for the past 10 years from EuroStat^2^. We propose that although total mortality may vary somewhat between calendar years, partly due to differing severity of seasonal epidemics such as influenza, the 10-year average in mortality is a solid baseline to compare against. The reported COVID-19 related mortality was retrieved from publicly available repositories (www.SCB.se and www.C19.se). In addition, we obtained data on number of COVID-19-related deaths from the regional morgue in Stockholm.

Reported COVID19-related deaths in Stockholm started in March 11^st^ (2020). Beginning of 2020 the weekly total mortality in the Stockholm region was slightly lower than the 10-year average mortality for the region, but there was a rapid increase in total mortality since COVID-19-related deaths were emerging in week 12 (Figure). Nevertheless, 24.9% of the excess mortality during the COVID-19 epidemic were not recognized as COVID-19-related, neither by public health data nor by the regional morgue.

**Figure.**
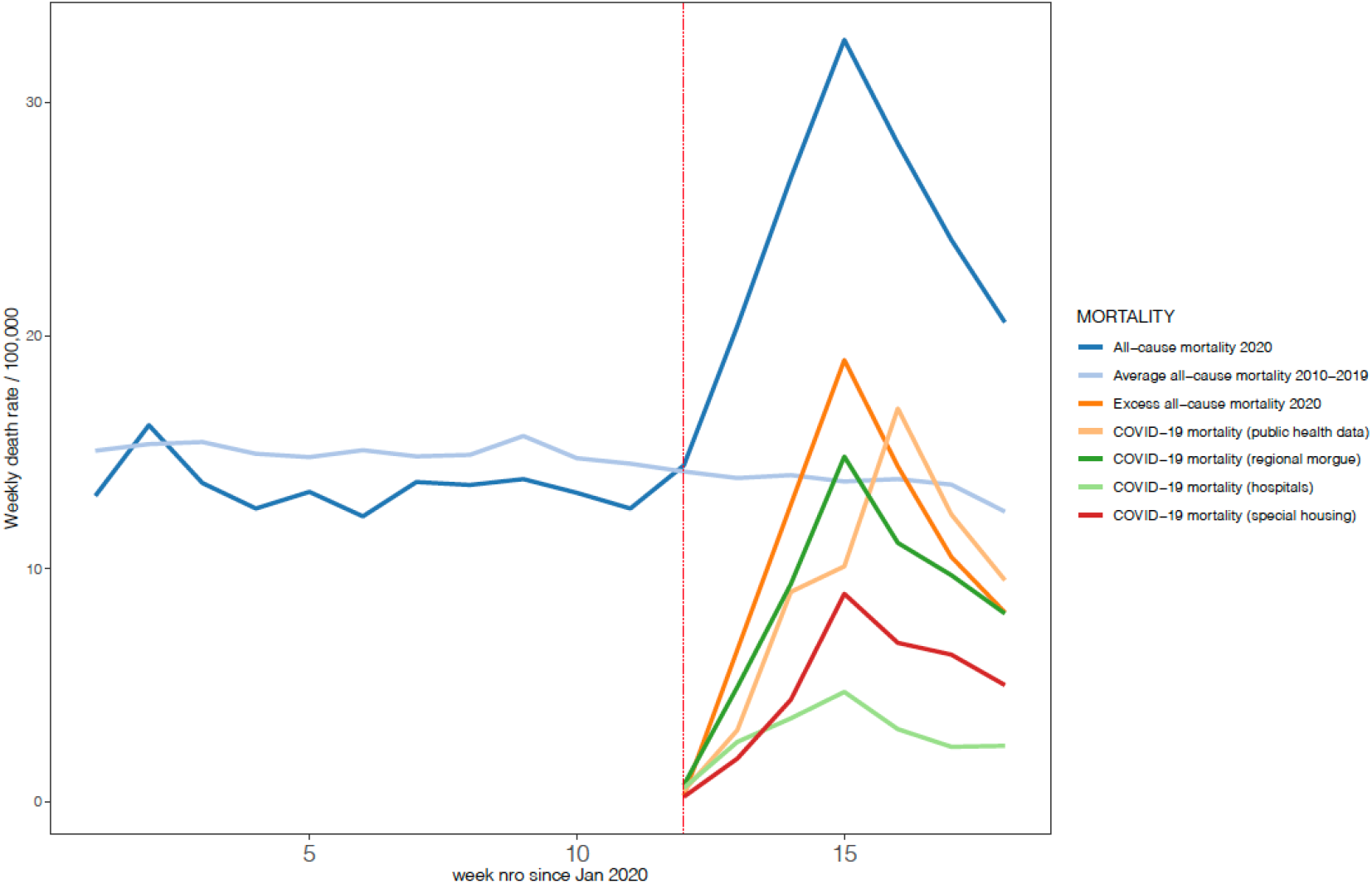
Excess mortality in 2020 in Stockholm between week 1 and 18. Total all-cause mortality in 2020 compared to the average all-cause mortality in the previous 10 years (2010-2019) and to the reported COVID-19-related mortality. The 2020 total all-cause mortality exceeds the average of the previous 10 years and becomes excess mortality at the same time as reporting of COVID-19-related deaths start emerging in week 12 (dashed vertical red line). Note that only a proportion of the excess mortality is recognized as COVID-19-related and that only a very small proportion is derived from recognized COVID-19-related deaths in hospitals.

The 2020 accumulated excess morality in the Stockholm region in week 18 was already +22.3% compared with the average over previous ten years. Reported COVID-19-related deaths in public health data, constituted 75.1% of the accumulated excess mortality in week 18 in Stockholm (Figure). Data from the regional morgue had similar findings, with minor shifts in the reported timing of deaths (Figure). In agreement with other countries^3^, only 23.6% of the reported COVID-19-related deaths had occurred in hospitals. Moreover, as expected, a much larger number of COVID-19-related deaths had occurred in other caretaking institutions, such as nursing homes for the elderly (Figure). For the remaining cases of reported COVID-19- related deaths the location of the death was not recorded on the death certificate.

In summary, we find that total excess mortality using average 10-year mortality as baseline, is well correlated to the reported COVID-19-related mortality in a region with multiple, population-based reporting systems. The well-established reporting infrastructure is likely to have identified a high proportion of all COVID-19-related deaths. The proportion of the total excess mortality that was recognized as COVID-19-related in our study (c. 75%) is probably a high estimate. That is, we predict that the total global death toll of the COVID-19 epidemic is conceivably more than 1.1 million deaths (>850,000/0.75). Strengths of our study is that we obtained public data from several sources, making the analysis of data robustness and reproducibility straightforward. Limitations include the fact that we focused on one region only and have no data on generalizability. Also, autopsies did not include systematic SARS-CoV-2 testing of the deceased which might have contributed to a number of unrecognized COVID-19-related deaths.

The Stockholm region has had a high COVID-19-related mortality^4^. In agreement with analysis from other countries ^5–8^, total excess mortality during the peak period seems to give a more comprehensive picture of the total burden of COVID-19-related deaths. Discrepancy between COVID-19-related mortality data and total excess mortality has also been noted by others^9,10^. Reporting systems based mostly on hospitals are likely to capture only a small fraction of the mortality burden. In our study, only 18% (0.24 × 0.75) of the excess mortality was reported as COVID-19-related based on hospital data. Some COVID-19 fatalities may have been unrecognized likely by their attributed comorbidities. The effects of the COVID-19 pandemic on the healthcare system and on healthcare-seeking behavior may also have caused some deaths, not directly induced by the virus.

Timely estimated mortality is essential to pinpoint the peak of mortality during epidemics, such as in COVID-19-related deaths. As such, all-cause total mortality has globally substantially increased during the COVID-19 pandemic. Taken together, we suggest that total excess mortality, calculated from ongoing all-cause mortality and compared to the all-cause mortality in the previous 10 years, during the epidemic peak period in a population may give a more comprehensive picture of the total burden of COVID19-related deaths.

## Data Availability

We retrieved mortality data for 2020 from the Swedish National Board of Health and Welfare and mortality estimates for the past 10 years from EuroStat2. We propose that although total mortality may vary somewhat between calendar years, partly due to differing severity of seasonal epidemics such as influenza, the 10-year average in mortality is a solid baseline to compare against. The reported COVID-19 related mortality was retrieved from publicly available repositories (www.SCB.se and www.C19.se). In addition, we obtained data on number of COVID-19-related deaths from the regional morgue in Stockholm.

https://appsso.eurostat.ec.europa.eu/nui

https://www.C19.se

